# Do Some Super-Spreaders Spread Better? Effects of individual heterogeneity in epidemiological traits

**DOI:** 10.1101/2022.04.19.22273976

**Authors:** Alexis S. Beagle, Sarah A. Budischak, Meggan E. Craft, Kristian Forbes, Richard Hall, David Nguyen, Clay E. Cressler

## Abstract

Many high-profile outbreaks are driven by super-spreading, including HIV, MERS, Ebola, and the SARS-Cov-2 pandemic. That super-spreading is a common feature of epidemics is immutable, however, the relative importance of 2super-spreaders to the outcome of an epidemic, and the individual-level traits that lead to super-spreading, is less clear. For example, an individual may contribute disproportionately to transmission by way of an extremely high contact rate or by way of low recovery, but how these two super-spreaders differ in their effect on epidemiological dynamics is unclear. Furthermore, epidemiological traits may often covary with one another in ways that promote or inhibit super-spreading. What patterns of covariation, and between what traits, are most likely to lead to large epidemics driven by super-spreading? Using stochastic individual-based simulations of an SIR epidemiological model, we explore how variation and covariation between transmission-related traits (contact rate and infectiousness) and duration-related traits (virulence and recovery) of infected individuals affects super-spreading and peak epidemic size. We show that covariation matters when contact rate and infectiousness covary: peak epidemic size is largest when they covary positively and smallest when they covary negatively. We did not see that more super-spreading always leads to larger epidemics, rather, we show that the relationship between super-spreading and peak epidemic size is dependent on which traits are covarying. This suggests that there may not necessarily be any general relationship between the frequency of super-spreading and the size of an epidemic.

## Introduction

Throughout the SARS-CoV-2 pandemic, the importance of super-spreading has been imprinted upon our cultural consciousness due to the high visibility of such cases and events; however, in the field of epidemiology, the significance of certain individuals being responsible for a disproportionate number of infections is par for the course. Not only have epidemiologists long known about the existence of super-spreaders, but they have shown that a significant number of high-profile outbreaks throughout modern history, including HIV, MERS, Ebola, and SARS, in addition to SARS-CoV-2, have also been driven by super-spreaders (Lui *et al*. 2020, Kain *et al*. 2021, May & Anderson, 1987, Wong *et al*. 2015, Gani & Leach 2004). In some well-studied outbreaks, such as SARS, information about identified super-spreaders has been recorded in great detail, going so far as to find their respective secondary infection rates, contact rates and other relevant measurements (Stein 2011, Shen *et al*. 2004). Comparing the characteristics of super-spreaders across outbreaks, however, reveals that super-spreading can be caused by variation (and potentially covariation) in many epidemiologically relevant traits. For example, one of the earliest and most quintessential examples of super-spreading, Typhoid Mary, was a super spreader by way of having low virulence and low recovery, while one of the documented SARS super-spreaders experienced high virulence and high contact rate (Brooks 1996, Shen *et al*. 2004). This limits the generality of conclusions regarding the role of super-spreaders on overall epidemiological dynamics: does it matter to epidemiological dynamics whether super-spreading arises through variation in transmission-related traits, such as contact rate or infectiousness (Vanderwaal and Ezenwa 2016, White *et al*. 2018, McCallum *et al*. 2017), versus through variation in duration-related traits, such as recovery rate or virulence (Gou and Jin 2017)? Moreover, is variation and covariation in some traits more likely to produce super-spreaders than (co)variation in others?

To illustrate, consider the pattern of higher infection prevalence and intensity in males compared to females observed for some types of parasites and some types of hosts (Muehlenbein & Bribiescas 2005, Luis *et al*. 2012, Vanderwaal & Ezenwa 2016, Kelly *et al*. 2018). These are thought to be due in part to elevated levels of testosterone (an immunosuppressant that likely prolongs infections; Zuk 2009, Foo *et al*. 2016) but also due to increased home range sizes which can lead to more infectious contacts (Klein *et al*. 2000, Luis *et al*. 2012). Here, individual variation in physiology and behavior act together to shape a potential pathway that might lead some males to become super-spreaders. Other examples include variation in attractiveness to vectors (Allan 2010, Takken & Verhulst 2013) or variation in disposition (being bold or shy) (in Siberian Chipmunks, *Tamias sibiricu;* Boyer *et al*. 2010; Vanderwall and Ezenwa 2016). How these different iterations of super-spreading compare to one another regarding influence on epidemiological dynamics is unclear.

We can use an epidemiological model to help identify traits that are likely to both impact epidemiological dynamics and lead to super-spreading. For example, an individual may become disproportionately important to spreading if it has a high transmission rate (which could be caused by a high contact rate or by high infectiousness) or if it is infectious for a long time (which could be caused by a very low recovery rate or mortality rate). Recently, authors have explored the potential importance of, not only variation, but covariation among individual traits to epidemiological dynamics as well (White *et al*. 2018, Gou & Jin 2017, Hawley *et al*. 2011). However, this was first suggested in the evolution of virulence literature (Cressler *et al*. 2016) which presupposes mechanistic links between disease parameters via within-host parasite replication. For instance, a trade-off (negative covariation) between virulence and transmission results when increasing within-host replication increases transmission by increased infectiousness/shedding but comes with a cost of increased virulence (Anderson & May 1982, Ewald 1983). There may also be a trade-off between virulence and recovery if high within-host replication reduces the ability of the immune system to clear the infection but also increases virulence (Anderson & May 1982, Alizon 2008). Lastly, a trade-off between contact rates and infectiousness can arise if high within-host replication increases infectiousness but also leads to increased sickness behavior that decreases host contact rates (Ewald 1994). These trade-offs make a strong case for covariation being possible in nature but are examples of negative covariation exclusively. However, there are plausible reasons to suspect that, in some cases, these correlations may be positive (Hawley *et al*. 2011). For example, Brookes and Ward (2019) recently showed that canines infected with rabies have increased contact rates due to disease-induced behavioral changes and increased shedding and thus increased infectiousness.

Empirical investigations show that super-spreading may arise through variation in traits related to both transmission (contact rate and shedding rate) and infection duration (virulence and recovery). Additionally, evidence of individual variation and clear rationale for covariation in epidemiologically relevant traits are well-documented in the literature. Despite this, the importance of individual variation/covariation on epidemiological dynamics, and how that relates to the way in which one becomes a super-spreader remains unclear. In an analysis of individual variation on outbreak dynamics, Lloyd-Smith *et al*. (2005) showed that increased variation is associated with more super-spreading events, but also increased probability of extinction, suggesting that more super-spreading does not necessarily imply larger epidemics. More recently, the effects of covariation on population-scale dynamics have been explored by several papers (Gou & Jin 2017, White *et al*. 2018, Hawley *et al*. 2011), but these have focused exclusively on covariation in transmission-related traits (i.e., susceptibility, contact rate, infectiousness). Here we use stochastic individual-based simulations of a simple SIR epidemiological model to explore how variation and covariation in transmission-related traits (contact rate and infectiousness) and duration-related traits (virulence and recovery) of infected individuals affects super-spreading, peak epidemic size and the subsequent relationship between super-spreading and peak epidemic size. We tested all directions of covariation (positive, negative, and none), with three levels of variation (low, moderate, and high) and three different basic reproductive numbers (R_0_; low, moderate, and high).

## Methods

### Model Description and Assumptions

In the absence of any variation, the epidemiological dynamics are determined by a simple SIR model (Fig. 1; Eq. 1-3):

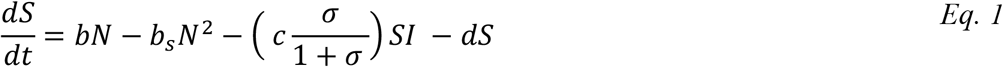

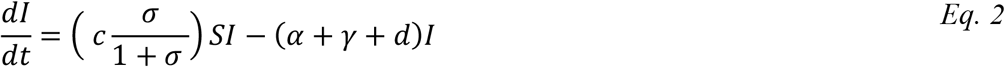

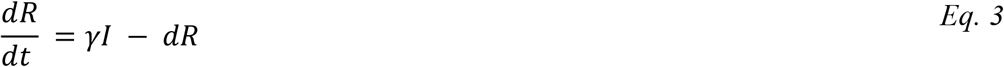

We assume that all individuals are born susceptible, and that birth rate is density-dependent (according to the parameter), with equal birth rates,, for all epidemiological classes. We decompose pathogen transmission rate into a contact rate process,, and an infection process (Fig. 1), with infectiousness determined by pathogen shedding rate () such that the probability of infection, given contact, is a saturating function of shedding rate. Infected hosts die due to infection at a per-capita rate, a, and recover into a permanently immune class at a per-capita rate γ. All hosts are assumed to die at the per-capita background rate, d.

**Figure 1.**
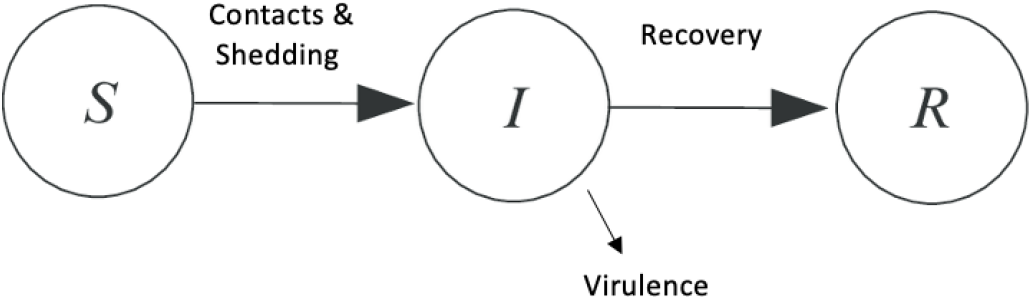
Flow diagram of SIR model. Contact and shedding determine movement from susceptible to infected, recovery determines movement from infected to recovered, and virulence determines infected individuals that do not recover. Births and deaths are not included in this diagram but are included in the SIR model used.

**Figure 1.**
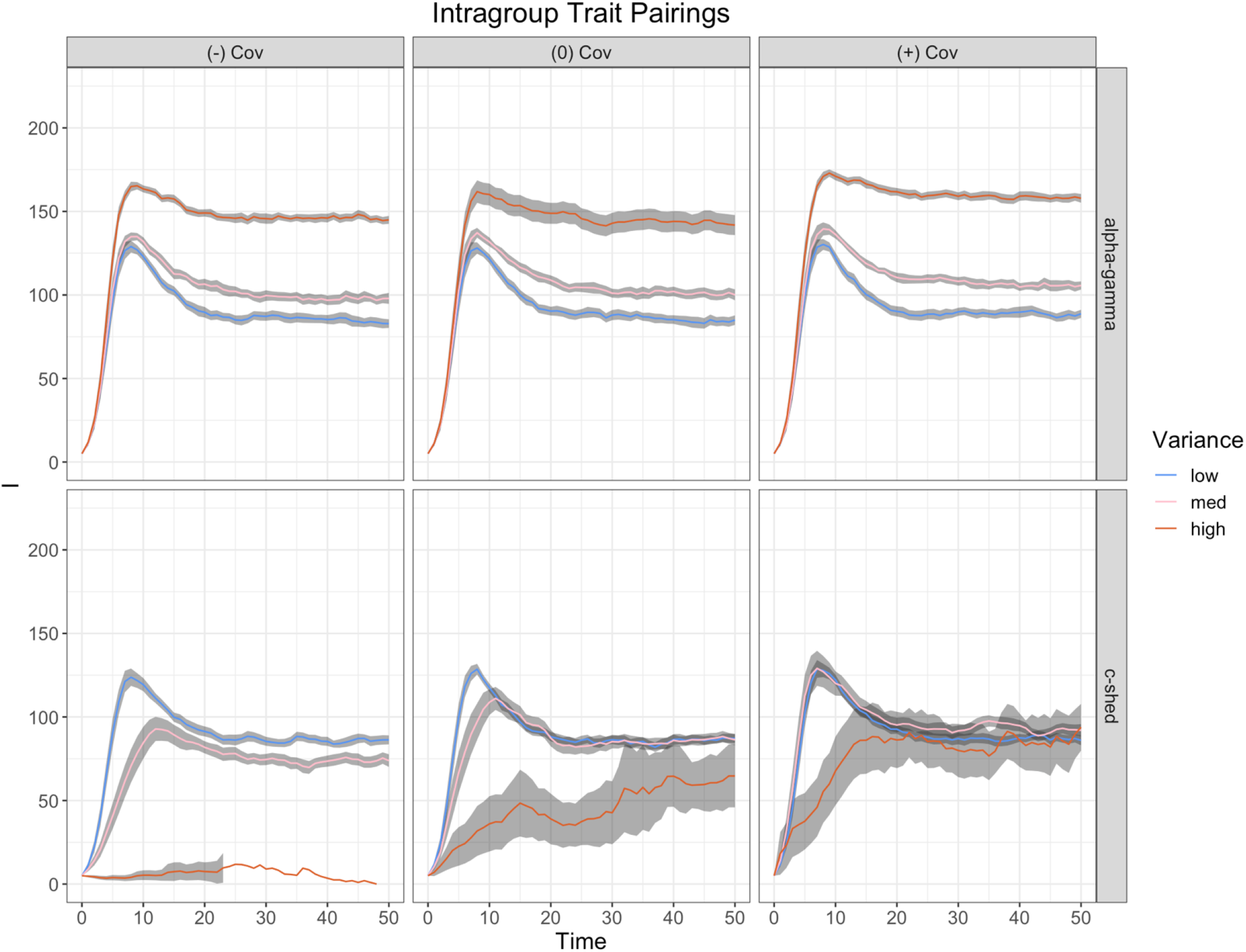
Dynamics of infected individuals over time for intragroup trait pairings at a moderate R_0_ for all levels of variation and directions of covariation. For each simulation, the initial number of susceptible hosts is given by the expected equilibrium population size in the absence of infection, minus the initial number of infected hosts, which was always 5.

To allow individuals to vary in contact rate, shedding rate, virulence, and recovery, we simulated the above SIR model with an individual-based model (IBM) built on the standard Gillespie algorithm for exact stochastic simulation of differential equations (Gillespie 1977). As in the standard algorithm, the probability of any of the possible events (birth, death, infection, and recovery) occurring at a moment in time is proportional to its rate, relative to all of the other events. Following DeLong (2016), we extend the algorithm by allowing each individual to have a unique set of parameters. Here we assume that epidemiological variation comes only through infected hosts, so that, for example, variation in contact rate is due to variation in the behavior of infected hosts, rather than susceptible hosts. The epidemiological traits of newly infected individuals are drawn from a multivariate normal distribution with fixed mean and covariance structure. With this individual-based model, we can track unique fates of each individual in the population, including the number of secondary infections each causes, while allowing population dynamics to emerge out of the stochastic processes of birth, death, infection and recovery.

To develop intuition about how (co)varation is likely to affect epidemiological dynamics, we can analyze how variation is predicted to affect the net reproductive rate, R_0_. For this simple model,

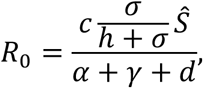

where *Ŝ* is the equilibrium number of susceptible hosts in the absence of infection, if one or more epidemiological parameters (co)vary, then R_0_ becomes a random variable and we can estimate its moments using a Taylor expansion. Table 1 gives the resulting expectations for R_0_, assuming variation and covariation between all epidemiological parameters; for full details of this derivation, see the appendix.

**Table 1:**
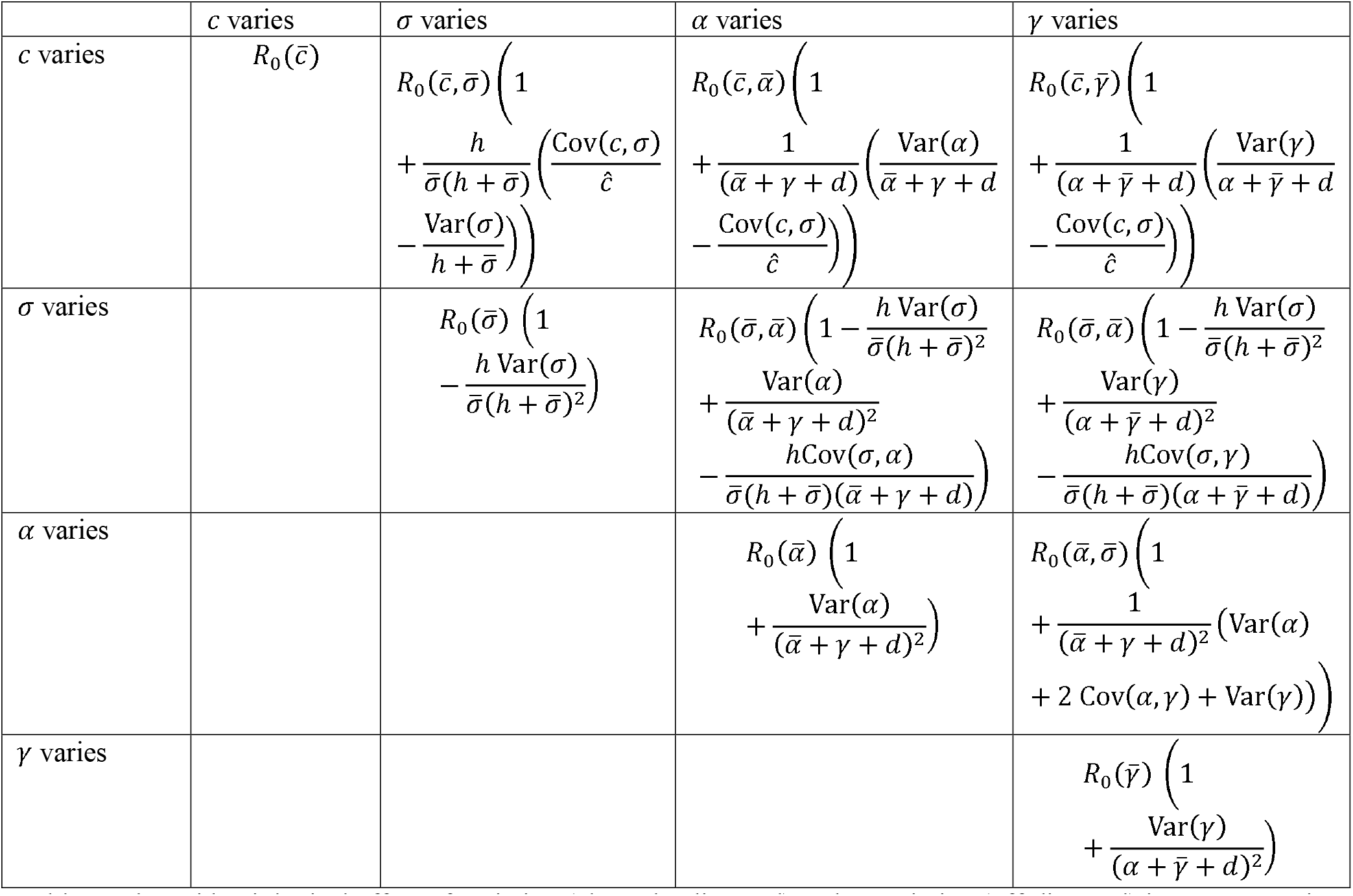
The epidemiological effect of variation (along the diagonal) and covariation (off-diagonal) in parameters is reported relative to *R*0 evaluated at the mean parameter values and 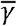.

| | $c$ varies | $\sigma$ varies | $\alpha$ varies | $\gamma$ varies |
| --- | --- | --- | --- | --- |
| $c$ varies | $R_0(\bar{c})$ | $R_0(\bar{c}, \bar{\sigma}) \left( 1 + \frac{h}{\bar{\sigma}(h + \bar{\sigma})} \left( \frac{\text{Cov}(c, \sigma)}{\hat{c}} - \frac{\text{Var}(\sigma)}{h + \bar{\sigma}} \right) \right)$ | $R_0(\bar{c}, \bar{\alpha}) \left( 1 + \frac{1}{(\bar{\alpha} + \gamma + d)} \left( \frac{\text{Var}(\alpha)}{\bar{\alpha} + \gamma + d} - \frac{\text{Cov}(c, \sigma)}{\hat{c}} \right) \right)$ | $R_0(\bar{c}, \bar{\gamma}) \left( 1 + \frac{1}{(\alpha + \bar{\gamma} + d)} \left( \frac{\text{Var}(\gamma)}{\alpha + \bar{\gamma} + d} - \frac{\text{Cov}(c, \sigma)}{\hat{c}} \right) \right)$ |
| $\sigma$ varies | | $R_0(\bar{\sigma}) \left( 1 - \frac{h \text{Var}(\sigma)}{\bar{\sigma}(h + \bar{\sigma})^2} \right)$ | $R_0(\bar{\sigma}, \bar{\alpha}) \left( 1 - \frac{h \text{Var}(\sigma)}{\bar{\sigma}(h + \bar{\sigma})^2} + \frac{\text{Var}(\alpha)}{(\bar{\alpha} + \gamma + d)^2} - \frac{h \text{Cov}(\sigma, \alpha)}{\bar{\sigma}(h + \bar{\sigma})(\bar{\alpha} + \gamma + d)} \right)$ | $R_0(\bar{\sigma}, \bar{\alpha}) \left( 1 - \frac{h \text{Var}(\sigma)}{\bar{\sigma}(h + \bar{\sigma})^2} + \frac{\text{Var}(\gamma)}{(\alpha + \bar{\gamma} + d)^2} - \frac{h \text{Cov}(\sigma, \gamma)}{\bar{\sigma}(h + \bar{\sigma})(\alpha + \bar{\gamma} + d)} \right)$ |
| $\alpha$ varies | | | $R_0(\bar{\alpha}) \left( 1 + \frac{\text{Var}(\alpha)}{(\bar{\alpha} + \gamma + d)^2} \right)$ | $R_0(\bar{\alpha}, \bar{\sigma}) \left( 1 + \frac{1}{(\bar{\alpha} + \gamma + d)^2} (\text{Var}(\alpha) + 2 \text{Cov}(\alpha, \gamma) + \text{Var}(\gamma)) \right)$ |
| $\gamma$ varies | | | | $R_0(\bar{\gamma}) \left( 1 + \frac{\text{Var}(\gamma)}{(\alpha + \bar{\gamma} + d)^2} \right)$ |

From this, we can make several predictions. In particular, variation in contact rate alone will have no impact on R_0_, whereas variation in either virulence or recovery will tend to increase R_0_. The effects of covariation are, of course, more complicated. If contact and shedding negatively covary (i.e., there is a trade-off between contact rate and infectiousness) then the expected R_0_ decreases. On the other hand, if contact rate negatively covaries with either virulence or recovery, then the expected R_0_ increases. If virulence and recovery positively covary, then the expected R_0_increases. In all other cases, it is impossible to make a general prediction, as the expected R_0_ depends on parameter values.

To confirm these predictions and more fully explore the effects of variation and covariation of transmission and duration-related traits on epidemiological dynamics, and, in particular, on the potential for super-spreading, we explored all possible combinations of contact rate (*c*), shedding rate (shed), virulence (alpha) and recovery (gamma), allowing parameters to covary with zero, positive, or negative covariation. We tested each model combination at three levels of variation (SD; 0.03, 0.15, 0.75) and three R_0_ values (R_0_; 1.5, 3.8, 8). The covariation was calculated from the correlation, set to either -0.5, 0, 0.5, and the variation. This resulted in a total of 54 model variants.

We carried out 50 stochastic simulations of each model, tracking the epidemiological dynamics and each infected individual’s trait values and number of secondary infections. We compared the peak epidemic size and the heterogeneity in the number of secondary infections for each model. To quantify heterogeneity, we use an estimate of the dispersion parameter (*k*) of a negative binomial model fit to the distribution of secondary infections for each simulation (Lloyd-Smith *et al*. 2005). We use the estimate of *k* as a measure of super-spreading: the larger the *k* value, the less heterogeneity there is in the number of secondary infections, and thus less super-spreading.

## Results

We found that our results were strongly determined by whether covarying parameters were duration (virulence, recovery) or transmission (contact rate, shedding rate) related. When trait pairings include one duration parameter and one transmission parameter (i.e., virulence and contact rate) we classify them as intergroup trait pairings. When trait pairings include parameters from the same group (i.e., virulence and recovery), we classify them as intragroup trait pairings. These groupings provide a general framework in which we can understand our results because, for most of our measures, results from simulations of intergroup trait pairings tend to be similar, whereas results from simulations of intragroup trait pairings are distinct to the trait pairing. We also found that when the simulated R_0_ is increased or decreased the general patterns are the same, but emphasized or deemphasized, respectively. Given this, our figures show only results from our simulations with a moderate R_0_ (see appendix for different R_0_ result figures). Similarly, when only one of the intergroup trait pairings is shown in our figures, it implies that the omitted simulation results are qualitatively identical to those that are shown (see appendix for all trait pairings).

### Effect of Variation on Peak Epidemic Size

When intergroup traits covary negatively (i.e., individuals with the largest recovery rate have the lowest shedding rate), regardless of which transmission and duration traits are covarying, increasing trait variation always increases the peak epidemic size (fig. 2&3). This agrees with the analytical results, which indicate that R_0_ is most likely to increase with variation if covariation is negative (Table 1). When traits vary independently or covary positively (i.e., there is a trade-off: individuals with the highest recovery rate have the highest shedding rate), the effect of increasing variation depends on which transmission traits are varying; in particular, if contact rate (c) varies, then increasing variation increases the peak with all directions of covariation; if shedding rate (shed) varies, however, increasing variation can increase, not affect, or even decrease the peak, depending on the direction of covariation (Fig. 3). For intragroup trait pairings, the effect of variation depends on the covarying traits (Fig. 3). When virulence (alpha) and recovery (gamma) covary, more variation always leads to a large epidemic peak regardless of covariation. This agrees with the analytical results (Table 1). When contact rate (c) and shedding rate (shed) vary, however, we see an increase in peak epidemic size when there is positive covariation, but no change in peak size with an increase in variation with no covariation. When these traits covary negatively, we see a decrease in peak epidemic size with an increase in variation. Interestingly, this does not agree with the analytical results, which indicate that increasing variation should always decrease R_0_ (Table 1).

**Figure 2.**
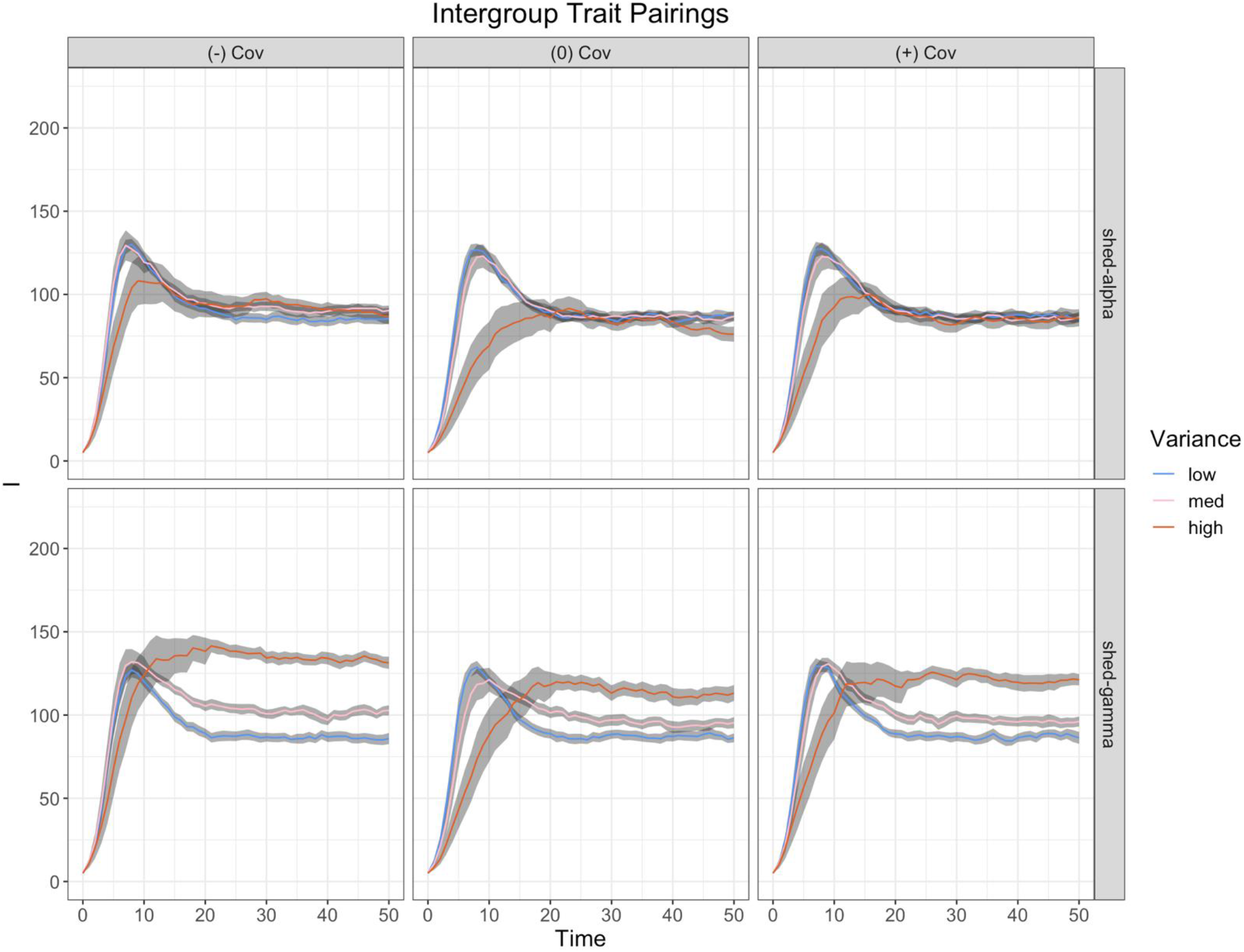
Dynamics of infected individuals over time for intergroup trait pairings at a moderate R_0_ for all levels of variation and directions of covariation. Note that the trait pairings of contact with virulence (alpha) and recovery (gamma) are omitted but can be found in the appendix.

**Figure 3.**
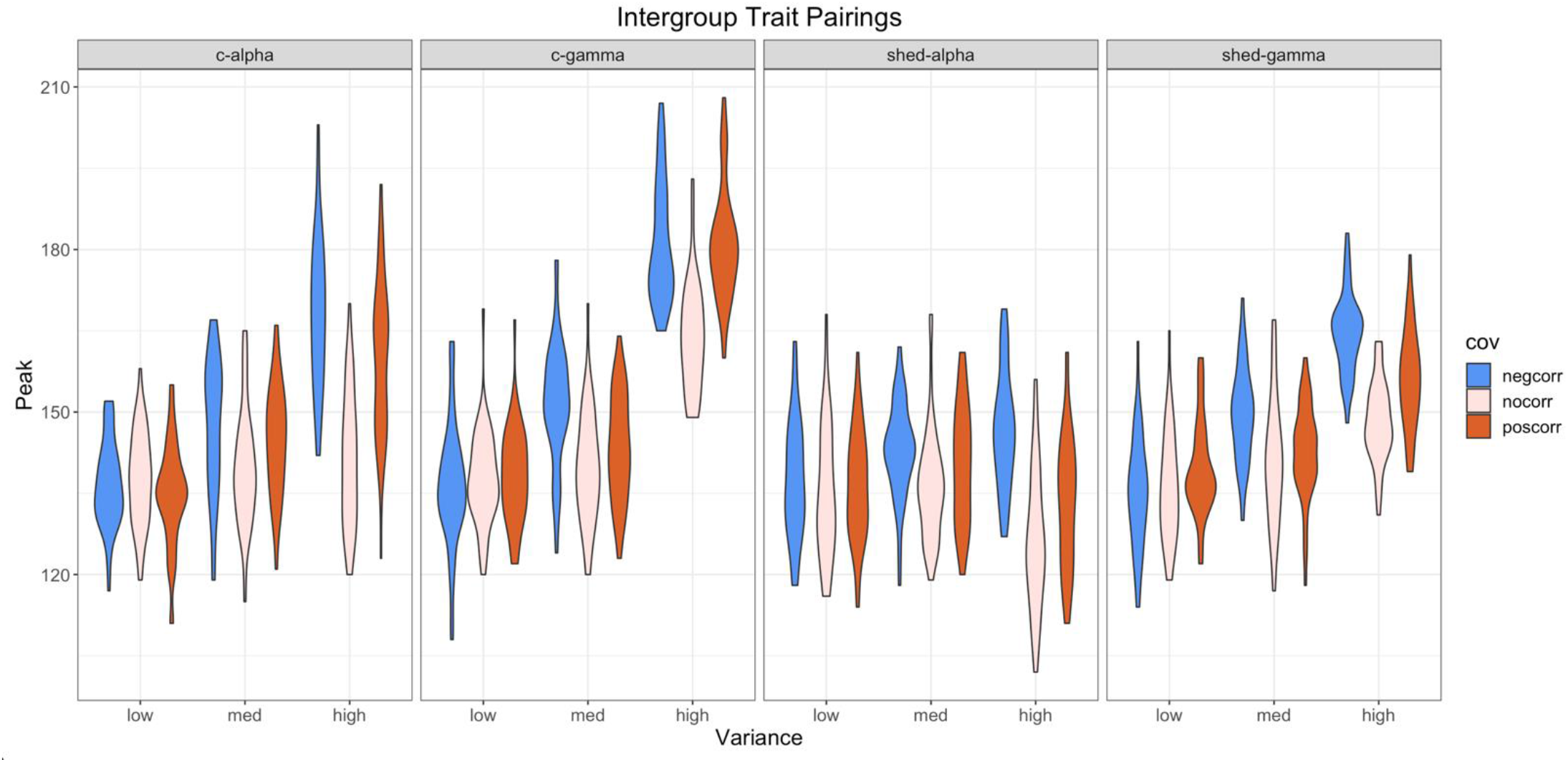
Distribution of peak epidemic sizes for intergroup trait pairings at all levels of variation and directions of covariation. Differences in violin width indicate the density of peak epidemic sizes at that value.

### Effect of Covariation on Peak Epidemic Size

We see that, at moderate variation, peak epidemic size of intergroup trait pairings is largest when covariation is negative, and larger when traits vary independently than when they covary positively (i.e., there is a trade-off) (fig. 3). At high variation, however, both negative and positive covariation result in larger peak epidemic sizes than independent covariation (fig. 3).

For intragroup trait pairings, we see the same pattern at all levels of variation: the largest peak sizes result from positive covariation and the smallest from negative covariation (fig. 3).

### Effect of Variation on Super-spreading (dispersion)

For all trait pairings, more variation leads to lower dispersion (k) which indicates more super-spreading (fig. 5). With negative covariation and the highest level of variation for our contact rate (c) and shedding rate (shed) simulations, however, no dispersion could be estimated due to extinctions.

### Effect of Covariation on Super-spreading (dispersion)

For intergroup trait pairings, negative covariation leads to the smallest dispersion values while positive leads to the largest (fig. 5). This indicates that negative covariation leads to more super-spreading than positive or independent covariation. For intragroup trait pairings, there is no clear pattern. There is no effect of covariation on super-spreading when virulence (alpha) and recovery (gamma) covary. When contact rate (c) and shedding rate (shed) vary we see that negative covariation leads to the largest dispersion values and positive covariation leads to the smallest.

This indicates that positive covariation causes more super-spreading than negative or independent covariation.

### Effect of super-spreading on peak epidemic size

We see that variation and covariation influence peak epidemic size and dispersion (super-spreading) (fig. 3&4); however, we do not find a direct interaction between dispersion and peak epidemic size (fig. 5). This indicates that the role of super-spreading during an epidemic is indirect and varied. For example, in the contact rate (c) and shedding rate (shed) panel of figure six, when looking at the moderate variation simulations (triangles), we see a trend of increasing peak epidemic size with a decrease in dispersion, but this trend is a function of the direction of covariation rather than only dispersion on peak epidemic size. Although we do not see a direct effect of super-spreading on epidemic size in our simulations, we do show under what conditions super-spreading should be an important driver of epidemiological dynamics. For example, when virulence (alpha) and recovery (gamma) vary, the effect of super-spreading on epidemiological dynamics should be negligible. In contrast, if intergroup traits are highly variable and covary negatively, we expect super-spreading to contribute to increasing the peak epidemic size because we see that despite approximately equal dispersion (k) for all directions of covariation at high variation, high dispersion with negative covariation leads to larger peak epidemic sizes.

**Figure 4.**
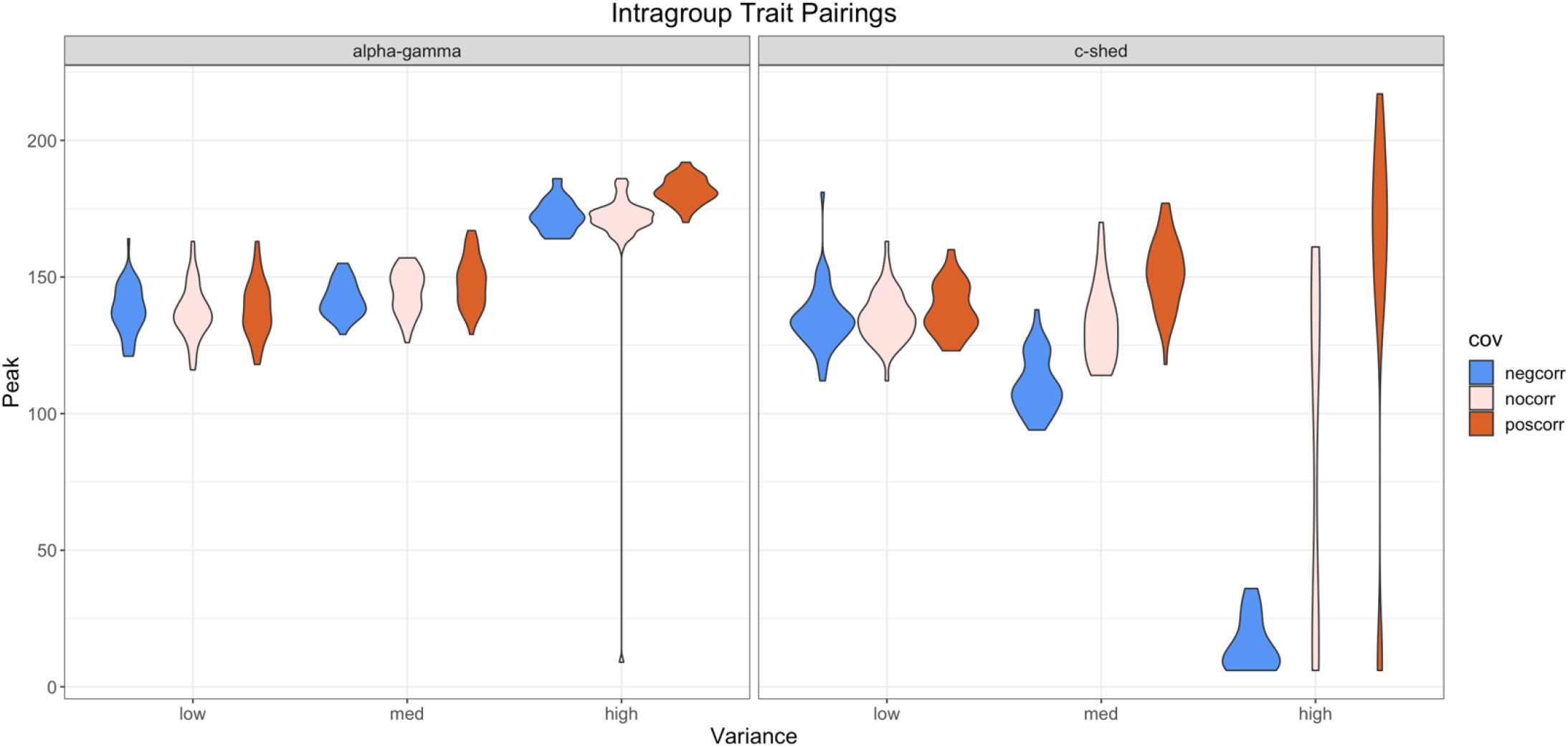
Distribution of peak epidemic sizes for intragroup trait pairings at all levels of variation and directions of covariation. Differences in violin width indicate the density of peak epidemic sizes at that value.

**Figure 4.**
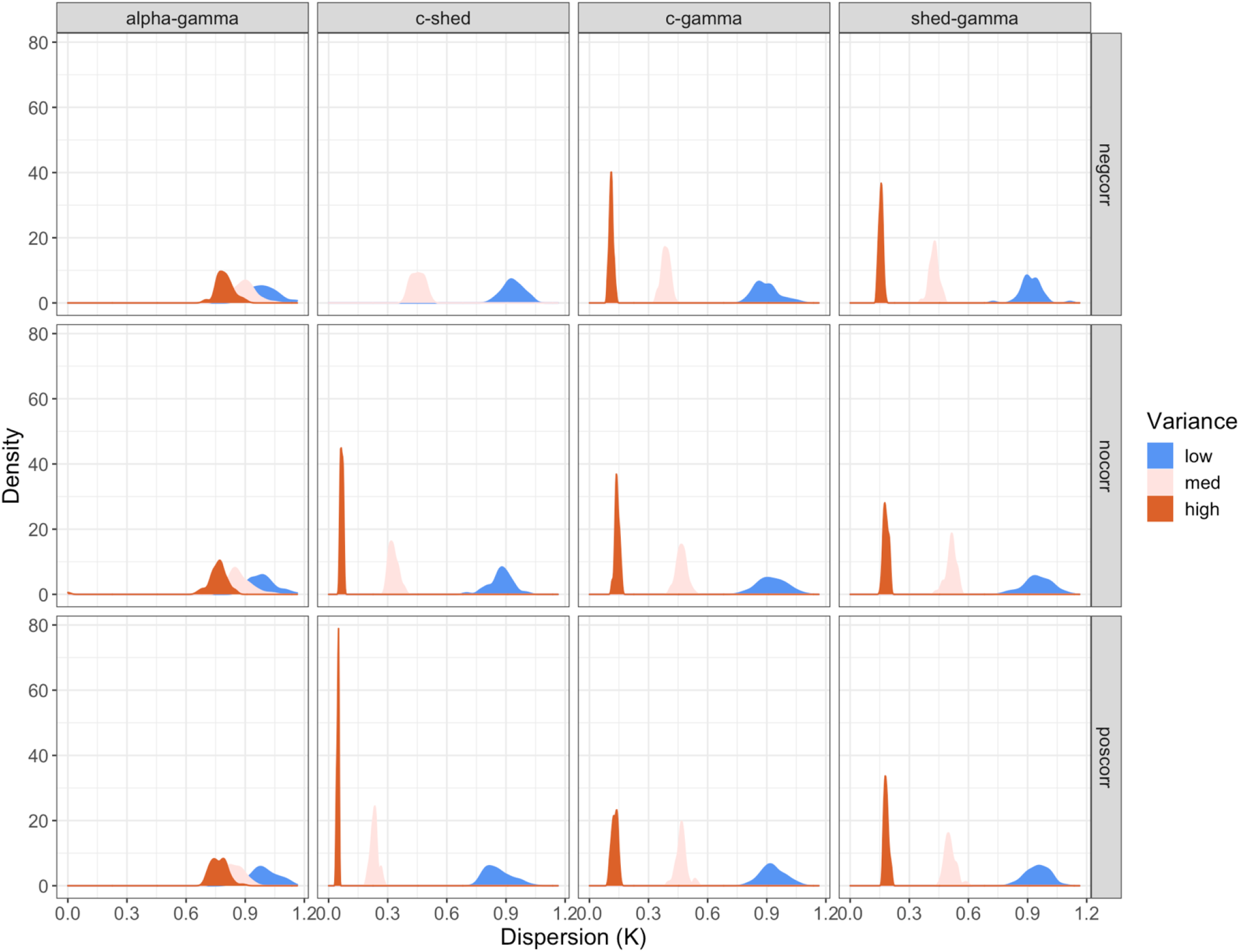
Distribution of dispersion (K) values for each trait pairing, level of variation and direction of covariation. Lower dispersion values indicate more super-spreading.

**Figure 5.**
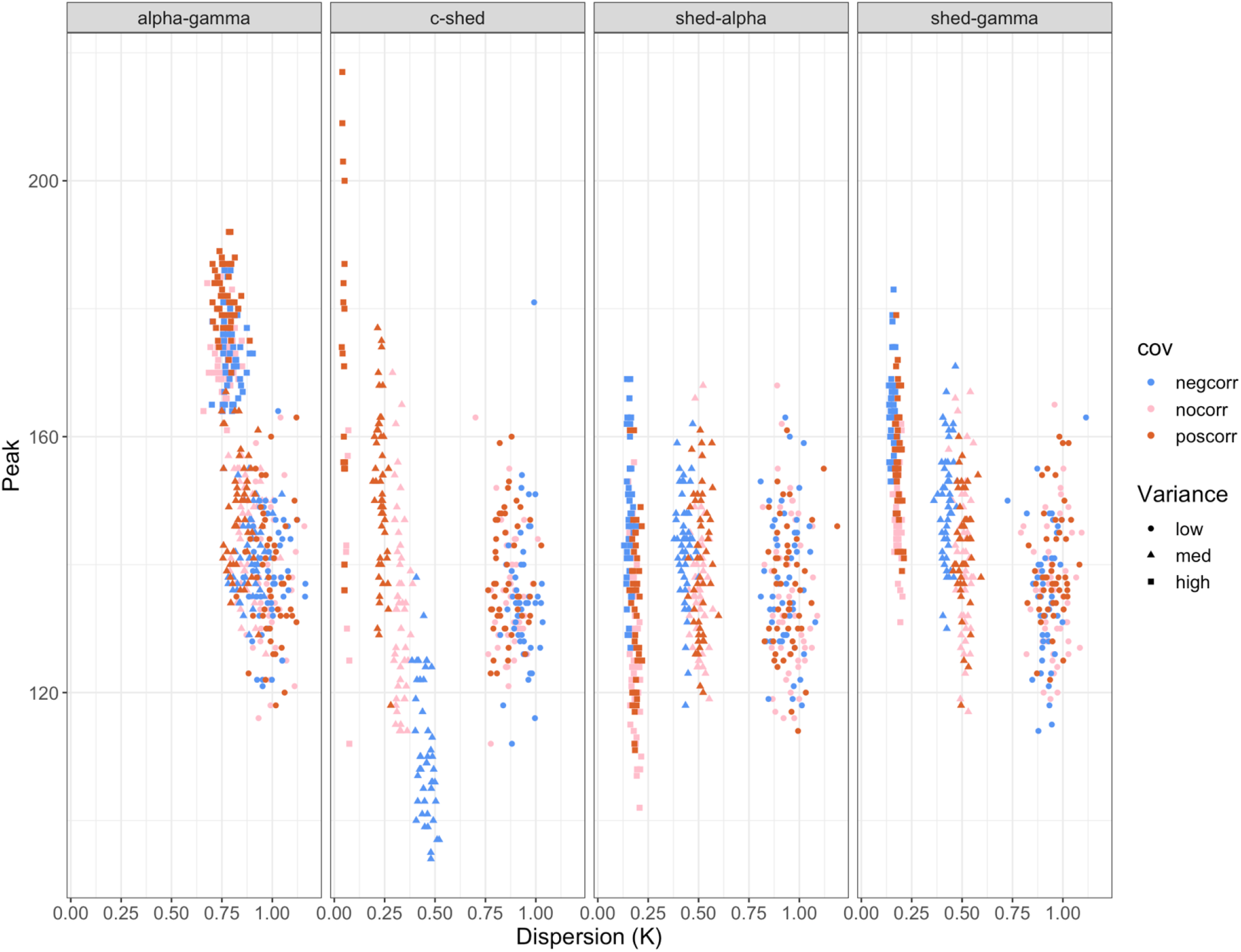
Relationship between dispersion (K) and peak epidemic size for all trait pairings, level of variance and direction of covariation.

## Discussion

In this study, we investigated how covariation in traits associated either with disease transmission (e.g., contact rate and infectiousness) or infection duration (e.g., recovery rate and virulence) influenced disease dynamics and individual heterogeneity in spreading. Previous investigations of this question have focused mainly on heterogeneity in traits associated with transmission and on epidemic dynamics, utilizing analytical approximation (Hawley *et al*. 2011), branching processes (Lloyd-Smith *et al*. 2005), and network-based approaches on static (Gou & Jin) and dynamic networks (White *et al*. 2018). In contrast, we built a stochastic, individual-based version of a classic SIR model with demography to understand how both epidemic and endemic dynamics are affected by heterogeneity (Fig. 1, 2). We also allowed for covariation between all combinations of transmission-and duration-related traits to investigate whether certain combinations of traits are more likely to give rise to larger epidemics or more super-spreading events.

Across simulations, we found that increasing variation almost always leads to an increase in the peak size of the epidemic (Fig. 3, 4). The only situation where this is less true is when contact and infectiousness covary, but this is driven by the fact that increasing variation increases the likelihood of epidemic fade-outs, as has been seen in other studies (Lloyd-Smith *et al*. 2005, White *et al*. 2018). When epidemics do take off in this case, they tend to be larger when there is more variability. Increasing variation also tends to increase individual heterogeneity in spreading, as evidenced by the decrease in the dispersion parameter, *k* (Fig. 5). The effect of variation is less pronounced when recovery and virulence covary – although there is increasing heterogeneity, the distributions of *k* are broadly overlapping suggesting that variation has less effect in this scenario. Interestingly, however, the observed values of *k* in our simulations are only comparable to existing empirical estimates in the high variation case. For example, Lloyd-Smith *et al*. 2005 estimated dispersion parameters to be less than 0.1 for all the pathogens they considered; this is in line with estimates for other pathogens (Melsew *et al*. 2019); we observed values of *k* that small only in the highest variation case, suggesting that there is considerable individual variability in epidemiological traits in most infectious disease systems.

The effects of covariation tended to be less pronounced, and to depend on the total amount of variation and whether the covarying traits affected transmission or duration (intragroup) or both transmission and duration (intergroup) (Fig. 3-5). For intergroup pairings, covariation only has an effect at the highest level of variation, and we observed larger epidemics and more super-spreading when traits covaried negatively (e.g., the most infectious individuals have the lowest recovery) or when they covaried positively (e.g., the most infectious individuals have the highest recovery). The negative covariation result is intuitive; why positive covariation (i.e., a trade-off) also leads to larger, more variable epidemics is less clear. For intragroup pairings, on the other hand, we see larger and more variable epidemics when covariation is positive (e.g., the most infectious individuals have the highest contact rates), as you might expect. Again, we see the largest effect of covariation sign when transmission traits covary, with epidemic fade-outs being much more likely under negative covariation than positive, especially at high variation.

Our observation that the effects of variation and covariation are largest when transmission traits covary, and smallest when duration traits covary, provides some validation for previous studies emphasis on these two aspects of the epidemiological process (Hawley *et al*. 2011, White *et al*. 2005). It is important to keep in mind that the expected R_0_ is identical across all trait pairings, and it is only the contact-infectiousness pairing that leads to frequent epidemic fade-outs, while also leading to the most super-spreading.

Another clear pattern that emerges from this study is that whether there is a relationship between super-spreading (measured by *k*) and the size of the epidemic, depends on the covarying traits. For example, when infectiousness and virulence covary (Fig. 5, shed-alpha), increasing variation increases super-spreading without any noticeable effect on peak size. However, when infectiousness and recovery covary (Fig. 5, shed-gamma), increasing variation increases both super-spreading and peak size. This suggests that there will not necessarily be any relationship between the frequency of super-spreading and the size of the epidemic, contrary to some previous work (Lloyd-Smith *et al*. 2005). Moreover, looking across stochastic simulations for a set of covarying traits and the sign and magnitude of covariation, we do not find that more super-spreading leads to larger peak.

In general, we find that it is critical to know which traits are covarying and how much they vary. Currently, there is little empirical evidence of covariation in nature. However, Hamilton *et al*. (2020) recently showed that in Tasmanian Devils (*Sarcophilus harrisii*) infected with devil facial tumor disease (DFTD), contact rates and infectiousness are negatively correlated: as the disease progresses and tumor surface area increases, Devils become more infectious (Obendorf & McGlashan 2008), but due to behavioral changes they have fewer contacts. Our simulations suggest that this negative covariation is likely important to the overall epidemiological dynamics given that, if these traits covaried positively we would expect larger epidemics and super-spreading, depending on the amount of individual variation. Much more common is empirical evidence of individual variation (VanderWaal & Ezenwa 2016, Ferrari *et al*. 2004, Melsew *et al*. 2019). For example, variation in pneumonia recovery rates have been documented in Bighorn sheep (*Ovis canadensis)* (Plowright *et al*. 2017). Our simulations show that, if recovery rate is covarying with another trait, epidemiological expectations may vary widely (Fig. 3). If recovery covaries with virulence (in any direction), we would expect minimal heterogeneity in number of secondary infections and only an increase in variation would lead to larger peak epidemic sizes (Fig. 3, 4). Conversely, if recovery covaries with contact rates, covariation (in any direction) would indicate an increase in peak epidemic size and super spreading being an important driver of epidemiological dynamics. The same expectations hold for individual heterogeneity in disease-induced mortality, which has been observed in a number of amphibian populations due to variation in parasite aggregation (Wilber *et al*. 2020).

Although documented cases of measured covariation are relatively rare, there is good reason to suspect that it is likely widespread and has the potential to largely influence population-scale disease dynamics. With this, our results highlight another nuance to understanding why certain individuals become super-spreaders. For example, an individual with a high contact rate may be considered a likely candidate for super-spreading, however, if contact rate covaries negatively with infectiousness (shedding rate), their influence on the epidemiological dynamics might be minimized, despite having many contacts. Conversely, if contact rate covaries in any direction with another trait, such as recovery, the high contact rate of an individual may lead such an individual to becoming a super-spreader in more cases since both positive and negative covariation increase peak epidemic size. Research to uncover when covariation between transmission and/or duration traits occurs will be important to furthering our ability to predict and manage epidemics and super-spreading.

## Data Availability

All data produced in the present study are available upon reasonable request to the author

## Reference

Alizon, S., Hurford, A., Mideo, N., & Baalen, M. V. (2009). Virulence evolution and the trade-off hypothesis: History, current state of affairs and the future. Journal of Evolutionary Biology, 22(2), 245–259. https://doi.org/10.1111/j.1420-9101.2008.01658.x

Anderson, R. M., & May, R. M. (n.d.). Coevolution of hosts and parasites. 16.

Barnett-Howell, Z., Watson, O. J., & Mobarak, A. M. (2021). The benefits and costs of social distancing in high-and low-income countries. Transactions of the Royal Society of Tropical Medicine and Hygiene, traa140. https://doi.org/10.1093/trstmh/traa140

Borremans, B., Reijniers, J., Hughes, N. K., Godfrey, S. S., Gryseels, S., Makundi, R. H., & Leirs, H. (2017). Nonlinear scaling of foraging contacts with rodent population density. Oikos, 126(6), 792–800. https://doi.org/10.1111/oik.03623

Brock, P. M., Murdock, C. C., & Martin, L. B. (2014). The History of Ecoimmunology and Its Integration with Disease Ecology. Integrative and Comparative Biology, 54(3), 353–362. https://doi.org/10.1093/icb/icu046

Brookes, V. J., Dürr, S., & Ward, M. P. (2019). Rabies-induced behavioural changes are key to rabies persistence in dog populations: Investigation using a network-based model. PLOS Neglected Tropical Diseases, 13(9), e0007739. https://doi.org/10.1371/journal.pntd.0007739

Brooks, J. (1996). The sad and tragic life of Typhoid Mary. CMAJ: Canadian Medical Association Journal, 154(6), 915–916.

Chowell, G., Castillo-Chavez, C., Fenimore, P. W., Kribs-Zaleta, C. M., Arriola, L., & Hyman, J. M. (2004). Model Parameters and Outbreak Control for SARS. Emerging Infectious Diseases, 10(7), 1258–1263. https://doi.org/10.3201/eid1007.030647

Cooper, L., Kang, S. Y., Bisanzio, D., Maxwell, K., Rodriguez-Barraquer, I., Greenhouse, B., Drakeley, C., Arinaitwe, E., G. Staedke S., Gething, P. W., Eckhoff, P., Reiner, R. C., Hay, S. I., Dorsey, G., Kamya, M. R., Lindsay, S. W., Grenfell, B. T., & Smith, D. L. (2019). Pareto rules for malaria super-spreaders and super-spreading. Nature Communications, 10(1), 3939. https://doi.org/10.1038/s41467-019-11861-y

DeLong, J. P., & Gibert, J. P. (2016). Gillespie eco-evolutionary models (GEMs) reveal the role of heritable trait variation in eco-evolutionary dynamics. Ecology and Evolution, 6(4), 935–945. https://doi.org/10.1002/ece3.1959

EBSCOhost | 122993920 | An Epidemiology Model of Devil Facial Tumor Disease in Tasmanian Devils. (n.d.). Retrieved March 24, 2022, from https://web.p.ebscohost.com/abstract?direct=true&profile=ehost&scope=site&authtype=crawler&jrnl=01973622&AN=122993920&h=k%2fH%2f1Ewne%2fKlKVc7VM53RFslrMFhBRP5ofDl%2bkkAj0iPZIbqnTDRGY8dr70SBPVklIOjopT6VGoVDbwP2Vaf0g%3d%3d&crl=c&resultNs=AdminWebAuth&resultLocal=ErrCrlNotAuth&crlhashurl=login.aspx%3fdirect%3dtrue%26profile%3dehost%26scope%3dsite%26authtype%3dcrawler%26jrnl%3d01973622%26AN%3d122993920

Estimation of the reproductive number of novel coronavirus (COVID-19) and the probable outbreak size on the Diamond Princess cruise ship: A data-driven analysis | Elsevier Enhanced Reader. (n.d.). https://doi.org/10.1016/j.ijid.2020.02.033

Ferrari, N., Cattadori, I. M., Nespereira, J., Rizzoli, A., & Hudson, P. J. (2004). The role of host sex in parasite dynamics: Field experiments on the yellow-necked mouse Apodemus flavicollis. Ecology Letters, 7(2), 88–94. https://doi.org/10.1046/j.1461-0248.2003.00552.x

Foo, Y. Z., Nakagawa, S., Rhodes, G., & Simmons, L. W. (2017). The effects of sex hormones on immune function: A meta-analysis. Biological Reviews, 92(1), 551–571. https://doi.org/10.1111/brv.12243

Gani, R., & Leach, S. (2004). Epidemiologic Determinants for Modeling Pneumonic Plague Outbreaks. Emerging Infectious Diseases, 10(4), 608–614. https://doi.org/10.3201/eid1004.030509

Gillespie, Daniel T. (1977). “Exact Stochastic Simulation of Coupled Chemical Reactions”. The Journal of Physical Chemistry. 81 (25): 2340–2361. CiteSeerX10.1.1.704.7634. doi:10.1021/j100540a008

Goldberg, T. L., Ruiz, M. O., Hamer, G. L., Brawn, J. D., Kitron, U. D., Hayes, D. B., Loss, S. R., & Walker, E. D. (2009). Host Selection by Culex pipiens Mosquitoes and West Nile Virus Amplification. The American Journal of Tropical Medicine and Hygiene, 80(2), 268–278. https://doi.org/10.4269/ajtmh.2009.80.268

Gou, W., & Jin, Z. (2017). How heterogeneous susceptibility and recovery rates affect the spread of epidemics on networks. Infectious Disease Modelling, 2(3), 353–367. https://doi.org/10.1016/j.idm.2017.07.001

Hamede, R. K., McCallum, H., & Jones, M. (2013). Biting injuries and transmission of Tasmanian devil facial tumour disease. Journal of Animal Ecology, 82(1), 182–190. https://doi.org/10.1111/j.1365-2656.2012.02025.x

Hamilton, D. G., Jones, M. E., Cameron, E. Z., Kerlin, D. H., McCallum, H., Storfer, A., Hohenlohe, P. A., & Hamede, R. K. (2020). Infectious disease and sickness behaviour: Tumour progression affects interaction patterns and social network structure in wild Tasmanian devils. Proceedings of the Royal Society B: Biological Sciences, 287(1940), 20202454. https://doi.org/10.1098/rspb.2020.2454

Handel, A., & Rohani, P. (2015). Crossing the scale from within-host infection dynamics to between-host transmission fitness: A discussion of current assumptions and knowledge. Philosophical Transactions of the Royal Society B: Biological Sciences, 370(1675), 20140302. https://doi.org/10.1098/rstb.2014.0302

Hawley, D. M., Etienne, R. S., Ezenwa, V. O., & Jolles, A. E. (2011). Does Animal Behavior Underlie Covariation Between Hosts’ Exposure to Infectious Agents and Susceptibility to Infection? Implications for Disease Dynamics. Integrative and Comparative Biology, 51(4), 528–539. https://doi.org/10.1093/icb/icr062

Hellriegel, B. (2001). Immunoepidemiology – bridging the gap between immunology and epidemiology. Trends in Parasitology, 17(2), 102–106. https://doi.org/10.1016/S1471-4922(00)01767-0

Kain, M. P., Childs, M. L., Becker, A. D., & Mordecai, E. A. (2021a). Chopping the tail: How preventing superspreading can help to maintain COVID-19 control. Epidemics, 34, 100430. https://doi.org/10.1016/j.epidem.2020.100430

Kawagoe, K., Rychnovsky, M., Chang, S., Huber, G., Li, L. M., Miller, J., Pnini, R., Veytsman, B., & Yllanes, D. (2021). Epidemic dynamics in inhomogeneous populations and the role of superspreaders. Physical Review Research, 3(3), 033283. https://doi.org/10.1103/PhysRevResearch.3.033283

Keeling, M. J., & Eames, K. T. D. (2005). Networks and epidemic models. Journal of The Royal Society Interface, 2(4), 295–307. https://doi.org/10.1098/rsif.2005.0051

Lau, M. S. Y., Dalziel, B. D., Funk, S., McClelland, A., Tiffany, A., Riley, S., Metcalf, C. J. E., & Grenfell, B. T. (2017). Spatial and temporal dynamics of superspreading events in the 2014–2015 West Africa Ebola epidemic. Proceedings of the National Academy of Sciences, 114(9), 2337–2342. https://doi.org/10.1073/pnas.1614595114

Lloyd-Smith, J. O., Schreiber, S. J., Kopp, P. E., & Getz, W. M. (2005). Superspreading and the effect of individual variation on disease emergence. Nature, 438(7066), 355–359. https://doi.org/10.1038/nature04153

Luis, A. D., Douglass, R. J., Hudson, P. J., Mills, J. N., & Bjørnstad, O. N. (2012). Sin Nombre hantavirus decreases survival of male deer mice. Oecologia, 169(2), 431–439. https://doi.org/10.1007/s00442-011-2219-2

Melsew, Y. A., Gambhir, M., Cheng, A. C., McBryde, E. S., Denholm, J. T., Tay, E. L., & Trauer, J. M. (2019). The role of super-spreading events in Mycobacterium tuberculosis transmission: Evidence from contact tracing. BMC Infectious Diseases, 19(1), 244. https://doi.org/10.1186/s12879-019-3870-1

Nyabadza, F., Chirove, F., Chukwu, C. W., & Visaya, M. V. (2020). Modelling the Potential Impact of Social Distancing on the COVID-19 Epidemic in South Africa. Computational and Mathematical Methods in Medicine, 2020, e5379278. https://doi.org/10.1155/2020/5379278

Obendorf, D. L., & Mcglashan, N. D. (n.d.). Critical reviews/Riviste critiche General topics/Argomenti generali.

Plowright, R. K., Manlove, K. R., Besser, T. E., Páez, D. J., Andrews, K. R., Matthews, P. E., Waits, L. P., Hudson, P. J., & Cassirer, E. F. (2017). Age-specific infectious period shapes dynamics of pneumonia in bighorn sheep. Ecology Letters, 20(10), 1325–1336. https://doi.org/10.1111/ele.12829

Ruiz, M., French, S. S., Demas, G. E., & Martins, E. P. (2010). Food supplementation and testosterone interact to influence reproductive behavior and immune function in Sceloporus graciosus. Hormones and Behavior, 57(2), 134–139. https://doi.org/10.1016/j.yhbeh.2009.09.019

Sexual dimorphism in immunity across animals: A meta□analysis. (n.d.). https://doi.org/10.1111/ele.13164

Shen, Z., Ning, F., Zhou, W., He, X., Lin, C., Chin, D. P., Zhu, Z., & Schuchat, A. (2004). Superspreading SARS Events, Beijing, 2003. Emerging Infectious Diseases, 10(2), 256–260. https://doi.org/10.3201/eid1002.030732

Siddle, H. V., Kreiss, A., Eldridge, M. D. B., Noonan, E., Clarke, C. J., Pyecroft, S., Woods, G. M., & Belov, K. (2007). Transmission of a fatal clonal tumor by biting occurs due to depleted MHC diversity in a threatened carnivorous marsupial. Proceedings of the National Academy of Sciences, 104(41), 16221–16226. https://doi.org/10.1073/pnas.0704580104

Stein, R. A. (2011). Super-spreaders in infectious diseases. International Journal of Infectious Diseases, 15(8), e510–e513. https://doi.org/10.1016/j.ijid.2010.06.020

University of St. Francis, Bruno, C. D., Comar, T., Benedictine University, Powell, M. O., University of St. Francis, Tameklo, A., & University of St. Francis. (2017). Age-Structured and Vaccination Models of Devil Facial Tumor Disease. SPORA: A Journal of Biomathematics, 3(1). https://doi.org/10.30707/SPORA3.1Bruno

VanderWaal, K. L., & Ezenwa, V. O. (2016). Heterogeneity in pathogen transmission: Mechanisms and methodology. Functional Ecology, 30(10), 1606–1622. https://doi.org/10.1111/1365-2435.12645

White, L. A., Forester, J. D., & Craft, M. E. (2018). Covariation between the physiological and behavioral components of pathogen transmission: Host heterogeneity determines epidemic outcomes. Oikos, 127(4), 538–552. https://doi.org/10.1111/oik.04527

Whittington, R., & Sergeant, E. (2001). Progress towards understanding the spread, detection and control of Mycobacterium avium subsp para-tuberculosis in animal populations. Australian Veterinary Journal, 79(4), 267–278. https://doi.org/10.1111/j.1751-0813.2001.tb11980.x

Wilber, M. Q., Briggs, C. J., & Johnson, P. T. J. (2020). Disease’s hidden death toll: Using parasite aggregation patterns to quantify landscapellevel host mortality in a wildlife system. Journal of Animal Ecology, 89(12), 2876–2887. https://doi.org/10.1111/1365-2656.13343

Wong, G., Liu, W., Liu, Y., Zhou, B., Bi, Y., & Gao, G. F. (2015). MERS, SARS, and Ebola: The Role of Super-Spreaders in Infectious Disease. Cell Host & Microbe, 18(4), 398–401. https://doi.org/10.1016/j.chom.2015.09.013

Woolhouse, M. E. J., Dye, C., Etard, J.-F., Smith, T., Charlwood, J. D., Garnett, G. P., Hagan, P., Hii, J. L.xK., Ndhlovu, P. D., Quinnell, R. J., Watts, C. H., Chandiwana, S. K., & Anderson, R. M. (1997). Heterogeneities in the transmission of infectious agents: Implications for the design of control programs. Proceedings of the National Academy of Sciences, 94(1), 338–342. https://doi.org/10.1073/pnas.94.1.338

Zhang, S., Diao, M., Yu, W., Pei, L., Lin, Z., & Chen, D. (2020). Estimation of the reproductive number of novel coronavirus (COVID-19) and the probable outbreak size on the Diamond Princess cruise ship: A data-driven analysis. International Journal of Infectious Diseases, 93, 201–204. https://doi.org/10.1016/j.ijid.2020.02.033

Zhang, Y., Jiang, B., Yuan, J., & Tao, Y. (2020). The impact of social distancing and epicenter lockdown on the COVID-19 epidemic in mainland China: A data-driven SEIQR model study [Preprint]. Epidemiology. https://doi.org/10.1101/2020.03.04.20031187

Zuk, M. (2009). The Sicker Sex. PLOS Pathogens, 5(1), e1000267. https://doi.org/10.1371/journal.ppat.1000267

